# Differentiating Borderline *HER2*-Expressing and *HER2*-Positive Cancers from Other Subtypes Using Serum Urokinase Plasminogen Activator

**DOI:** 10.64898/2026.01.15.26344197

**Authors:** Michael E. J. López Mujica, Suchanat Boonkaew, Nana Louise Christensen, Mette Abildgaard Pedersen, Kit Riegels Jørgensen, Mikkel H. Vendelbo, Elena E. Ferapontova

## Abstract

**Background:** *HER2*-positive (*HER2*+) cancers are associated with aggressive tumor development but also high response rates to targeted blockade treatments of the HER-2/*neu* signaling pathway leading to improved clinical outcome for the patient. Current clinical analysis of the *HER2* status primarily relies on solid tumor biopsies low-suitable for continuous real-time monitoring needed for possible adjustment of the treatment, while serum tests targeting blood-circulating HER-2/*neu* fragments often show conflicting tumor-serum relations.

**Methods:** A cellulase-linked aptamer sandwich assay was used for detection of total urokinase plasminogen activator (uPA) and its different forms in serum of cancer patients and healthy individuals. Serum uPA levels were correlated with solid biopsy results and relevant clinical data extracted from electronic patient records, and FDG-PET/CT scanning.

**Results:** We show that serum uPA allows precise stratification of patients with *HER2*+ cancers and cancers with *HER2* borderline expression. Serum levels of total uPA 96.6% accurately informed about *HER2*+ tumor status in a cohort of 85 patients, with a *HER2*+ cut-off value of 0.976 ng mL^-1^.

**Conclusions:** The established liquid biopsy test for serum uPA has potential for accurate diagnosis and staging of patients with *HER2*+ cancers and “borderline” cancers requiring further confirmatory (or rejection) testing.

## 1. Background

Despite huge advancements in treatment, cancer remains the second leading cause of death, with almost 10 million fatalities reported in 2020 worldwide [1]. Aggressive forms of breast, esophageal, lung, liver, and pancreatic cancer have particularly poor prognoses and contribute significantly to the death toll, showing five-year survival rates below 20% [2]. To improve treatment outcomes, more advanced precision medicine approaches are needed, including diagnostic tools for early cancer detection tailored to individual cancers that would ease fast decision making on which treatment patients will benefit from.

One of the most promising analytical tools, both for early cancer detection and its continuous treatment monitoring, is liquid biopsy [3] defined as a laboratory testing of a sample of body fluids (blood, urine, etc.) for biomolecules/cancer cells released by tumor. By providing non-invasive and easy access to specific tumor biomarkers through a simple blood draw, liquid biopsy is not biased by selection of the tumor region as solid biopsy. Yet, its largest challenge is finding robust liquid-biopsy molecular biomarkers of specific cancers such as tumor-specific proteins currently defined and detected largely by tissue biopsy [4–6].

Human Epidermal growth factor Receptor-2 (HER-2/*neu*), a glycoprotein complex belonging to the family of receptor tyrosine kinases, refers to such tumor-specific protein biomarkers when overexpressed by tumor cells in a number of aggressive cancers, such as breast, lung, colorectal, bladder, and gastro-esophageal cancers [7,8]. *HER2*-positive tumors (showing HER-2/*neu* immunohistochemistry (IHC) score 3+) tend to exhibit aggressive growth and high potential for metastasis associated with poor clinical outcomes [7]. In gastroesophageal cancer patients, *HER2*-positive tumors require targeted blockade treatments of the HER-2/*neu* signaling pathway [9]. The same refers to from 15% to 20% of breast cancers, where *HER2* overexpression correlates with a particularly aggressive tumor development, making HER-2/*neu* targeted therapies important for improving patient prognosis [8,10]. Concurrently, score 1+/2+ tumors with no *HER2* gene amplification (*HER2*-low breast cancers) also responded well to *HER2*-targeted therapies (the DESTINY-Breast04 trial) [11], which is poorly understood but, in terms of treatment, challenges current classification of *HER2*-associated cancers. Accurate monitoring of the *HER2* tumor status over disease progression is thus key to assessing the response and adjusting the treatment. Yet, current clinical analysis of *HER2* cancer subtypes primarily depends on solid biopsies, providing tissue for detecting expression of the protein itself by IHC, supported by examination of possible amplification of the *HER2* gene through fluorescence in situ hybridization (FISH) [12,13]. Both, approved by FDA, are inconvenient to the patient, as multiple biopsies should be taken.

Liquid biopsy analysis of the extracellular domain of HER-2/*neu*, proteolytically cleaved and released into the bloodstream by tumor cells, was suggested as alternative to solid tumor HER-2/*neu* analysis [12,13], enhancing diagnostic accuracy [14,15]. Yet, reports on HER-2/*neu* serum diagnostics are inconsistent and show conflicting tumor-serum correlations [13,15–22].

Further evidences occur that urokinase plasminogen activator (uPA), a serine protease, implicated in the processes of tumor invasion and metastatic spread [23,24], is overexpressed in *HER2*-positive tumors [25,26], with both *HER2* and uPA receptor (uPAR) genes co-amplified most frequently in the same patient’s cancer tissue extracts [26]. uPA is a part of the blood fibrinolytic system inducing peri-cellular proteolysis either by degrading extracellular-matrix (ECM) components or by activating latent proteases or growth factors [27]. Along with its receptor, uPAR, uPA not only triggers a cascade of proteolytic events occurring during tumor invasion and metastatic spread but also plays a larger role in cancer development, from tumorigenesis to metastasis [24,28]. Hitherto, most studies have focused on evaluation of uPA as a prognostic biomarker of disease-free and overall survival rates [29]. Its expression in tumor cells was correlated with survival rates in node-negative breast cancers [30–32], primary invasive breast cancers [33], primary breast cancers [34], and primary colorectal cancers [35]. Overexpression of uPAR in *HER2*-positive breast tumors was shown to contribute to aggressive metastatic phenotype of cancer with poor clinical outcome [25], and elevated serum uPA in cancers patients, above 2.5 ng mL^-1^, was reported as a cut-off for metastatic vs. non-metastatic breast cancer type and also predicating shorter survival rates [36].

Given that uPA is overexpressed in *HER2*-positive breast tumors [25,26], we hypothesized that serum uPA might be a consistent liquid biopsy biomarker of *HER2*-positive cancers. Here, we assessed serum uPA in 15 healthy volunteers and 85 cancer patients diagnosed with mostly breast and esophagus cancers of different phenotypes. For serum analysis, we used analytically validated electrochemical cellulase-linked aptamer-sorbent assay (e-ELASA) on magnetic beads [37–39], earlier shown to be an accurate tool for new biomarker’s discovery [39] (**Scheme 1**). We show that serum uPA informs correctly about *HER2*-positive tumors and appears to be an accurate biomarker of this cancer phenotype.

**Scheme 1.**
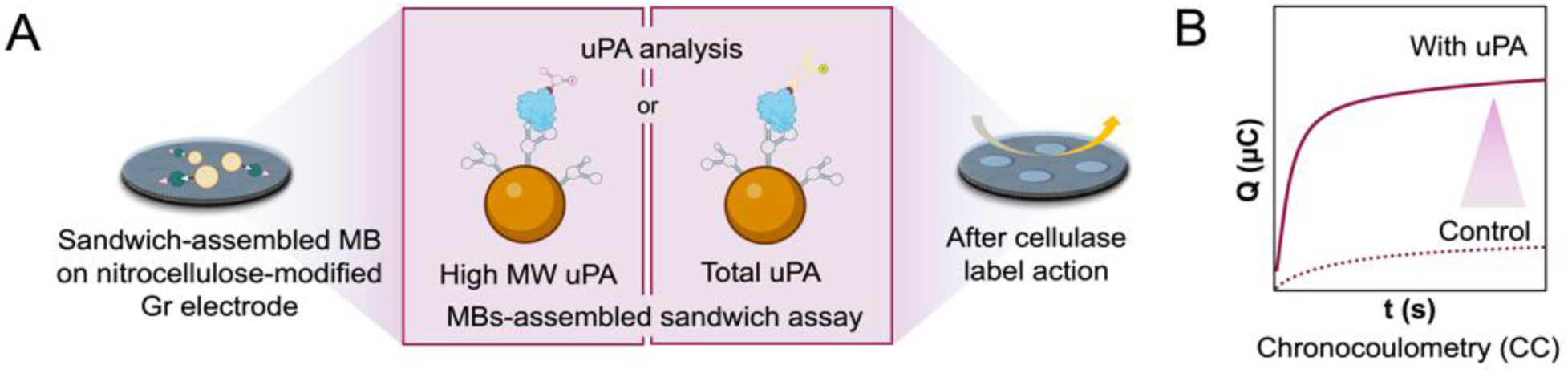
(**A**) Principle of the uPA detection by the sandwich aptamer1-uPA-aptamer2-cellulase assay on magnetic beads (MBs). The protein binds to the aptamer1-modified MBs, then the cellulase-conjugated reporter aptamer2 binds to uPA, with this completing the sandwich assembly. When applied onto nitrocellulose-modified electrodes, cellulase digests the film, changing its electrical properties, which are (**B**) then read out chronocoulometrically.

## 2. Methods

### Materials and reagents

Urokinase (uPA, 411 amino acids; MW: 54 kDa, a two-chain glycoprotein isolated from human urine, specific activity: 187,973 IU mg^-1^) was delivered by ProSpec (Israel) as a sterile filtered lyophilized powder. Aptamers specific for uPA [40] ordered from Metabion (Germany) were: uPA02 as a capture aptamer for both high molecular weight (HMW uPA) and total uPA (42-mer: 5’-biotin-CAA GCG GGG GTG AGA GAT CTG TCA GTA CGA GCT GGG TTT GCG-3’); uPA08, as a reporter aptamer solely for HMW uPA (41-mer: 5’ amine-C_6_-CAG CGG TAG GGG TTA TAT AGC TGC GCC ATA GGG TAC TCG TG-3’); and uPA21, as a reporter aptamer for total uPA (82-mer: 5’ amine-C_6_-AGG TAG AGG AGC AAG CCA TCG GAG GTA CTC ACC GAC GCT GAA CTC CAT AGA ATG TGG TGA TGG ATG CGT GAT CGA ACC TAC C-3’). All stock solutions (uPA, aptamers, and serum samples) were prepared/diluted with a 10 mM phosphate buffer solution containing 150 mM NaCl, pH 7.4 (PBS), at room temperature, rt (20 ± 2°C), stored at −20°C until used. Additional details on reagents are in Supporting Information, **SI**.

### Sandwich aptamer assay

Protocols for electrode handling, streptavidin-modified magnetic beads (MB) functionalization, and cellulase conjugation to reporter aptamers are given in SI. For calibration curves construction, 40 μL of uPA02-modified MBs were mixed with 960 μL of uPA solutions (from 10 aM to 100 pM) either in PBS, pH 7.4, or in 10% serum, and incubated for 30 min at rt with 300 rpm shaking. uPA-spiked serum samples were prepared with human serum from male AB plasma diluted 10 times with PBS. MBs were then washed 3×1 mL with 0.1% BSA in PBS, pH 7.4, the BSA solution being decanted. Next, 100 μL of 1 μM cellulase-aptamer bioconjugate (uPA08 or uPA21) were added to MBs and allowed to react for 30 min at rt under 300 rpm shaking, then being washed 3×100 μL with 0.1% BSA in PBS. After decanting the BSA solution, MBs with fully assembled sandwiches were resuspended in 200 μL of PBS, pH 5, and 5 μL of sandwich suspension were dropped onto the nitrocellulose-modified electrodes and incubated for 20 min at rt. Finally, MBs were water-rinsed from the electrode surface, and the electrodes were chronocoulometrically (CC) tested, at 0.3 V (*t*-interval 0.1 s; *t*-run 10 s). The control (blank) experiments were performed by running the MB-assembly assay without uPA or serum uPA, but only in PBS or serum, with the same electrodes further exposed, after being washed with water, to sandwiches assembled on MBs in sample solutions. The change in the charge Δ*Q* was calculated by subtracting the blank response (*Q*_0_) from the response of uPA-containing samples (*Q*_uPA_). All analytical detections, also with patients’ samples, were performed in triplicates, each time with a new electrode.

### Sample preparation and statistical analysis

Serum samples were prepared as previously described [39]. Briefly, the collected blood samples were centrifuged at 2000×g for 10 min to remove clots, and the resulting supernatant (serum) was then stored at −20°C until use. All serum samples were diluted 10-fold with PBS and then assayed as described above. Cancer biopsy results and other relevant clinical data (pathology reports and biochemical laboratory results) were extracted from the electronic patient records. All details on statistical analysis of the data are in SI.

## 3. Results

### 3.1. Analysis of total serum uPA

First, cellulase-linked e-ELASA, developed and applied earlier for accurate detection of serum HER-2/*neu* and PSA [37–39,41], was adapted for analysis of serum uPA by using a couple of uPA02 (capture) and uPA21 (reporter) aptamers specific for total uPA (**Scheme 1**). These were DNA aptamers, inexpensive compared to the previously used fluorinated RNA aptamer [42] yet similarly stable in serum. Calibration curves were constructed and used for uPA analysis in human samples (**Figure S1A, S1B, S2**, **SI**). Sensitivity of total uPA detection was (20.28 ± 5.21) µC fM^-1^ in PBS and (19.14 ± 4.50) µC fM^-1^ in serum (relative difference, RD=6.0%), and the limit of detection (LOD) determined by IUPAC as “*the smallest concentration of analyte in the sample that can be reliably distinguished from zero*” was 1 aM, both in serum and PBS (0.430 aM when calculated as 3×*σ*/*S*, where *σ* is the standard deviation of the blank signal and *S* is the sensitivity of analysis (the slope of the calibration curve).

Serum uPA was then analyzed in 15 healthy individuals and 85 cancer patients (54 breast cancer, 25 esophagus cancer, 5 gastric cancer and 1 cardia cancer), of whose 80 patients were diagnosed with different degrees of *HER2* gene expression (normal expression correlates with IHC HER-2/*neu* score 0 and 1+, borderline expression – with score 2+, and overexpression – with score 3+ indicating *HER2*-positive cancers [43]), including 18 patients with a borderline expression (2 of those with confirmed *HER2*-positive breast cancer) and 19 patients with an overexpressed *HER2* gene. Here, for convenience, this 37 patients’ group is also referred to as “allegedly *HER2*-positive”, yet corresponding to scores 2+ and 3+, of which only score 3+ indicated *HER2*-positive cancers, while score 2+ patients needed additional testing of the *HER2* gene amplification degree, to either confirm or reject the *HER2*-positive diagnosis. Further, we provide separate analytical results for the *HER2* borderline expression and overexpression cases. For 5 cancer patients their *HER2* status was not identified and thus considered as not *HER2*-positive, yet they were excluded from final analysis (**Figure S3, SI**). Serum uPA levels were increased statistically significantly in the allegedly *HER2*-positive 37 patients’ group (a 95% confidence interval), compared to the uPA levels in cancer patients with low and normal expression of *HER2* gene and healthy individuals (**Figure 1A, 1B**). The average concentration of total uPA in allegedly *HER2*-positive cases reached (4.030±2.611) ng mL^-1^ (*p* < 0.0001), 200-fold exceeding uPA levels detected in the serum of other cancer patients, (0.020±0.013) ng mL^-1^ (*p* < 0.001), and 444-fold exceeding serum uPA in a healthy cohort, (0.009±0.006) ng mL^-1^ (*p* < 0.001). Only 1 of 37 analyses was visibly out of range, leading to 96.6% accuracy of the *HER2* borderline expression and overexpression detection by serum uPA, indicating accurately the extent of HER-2/*neu* expression in both cases.

**Figure 1.**
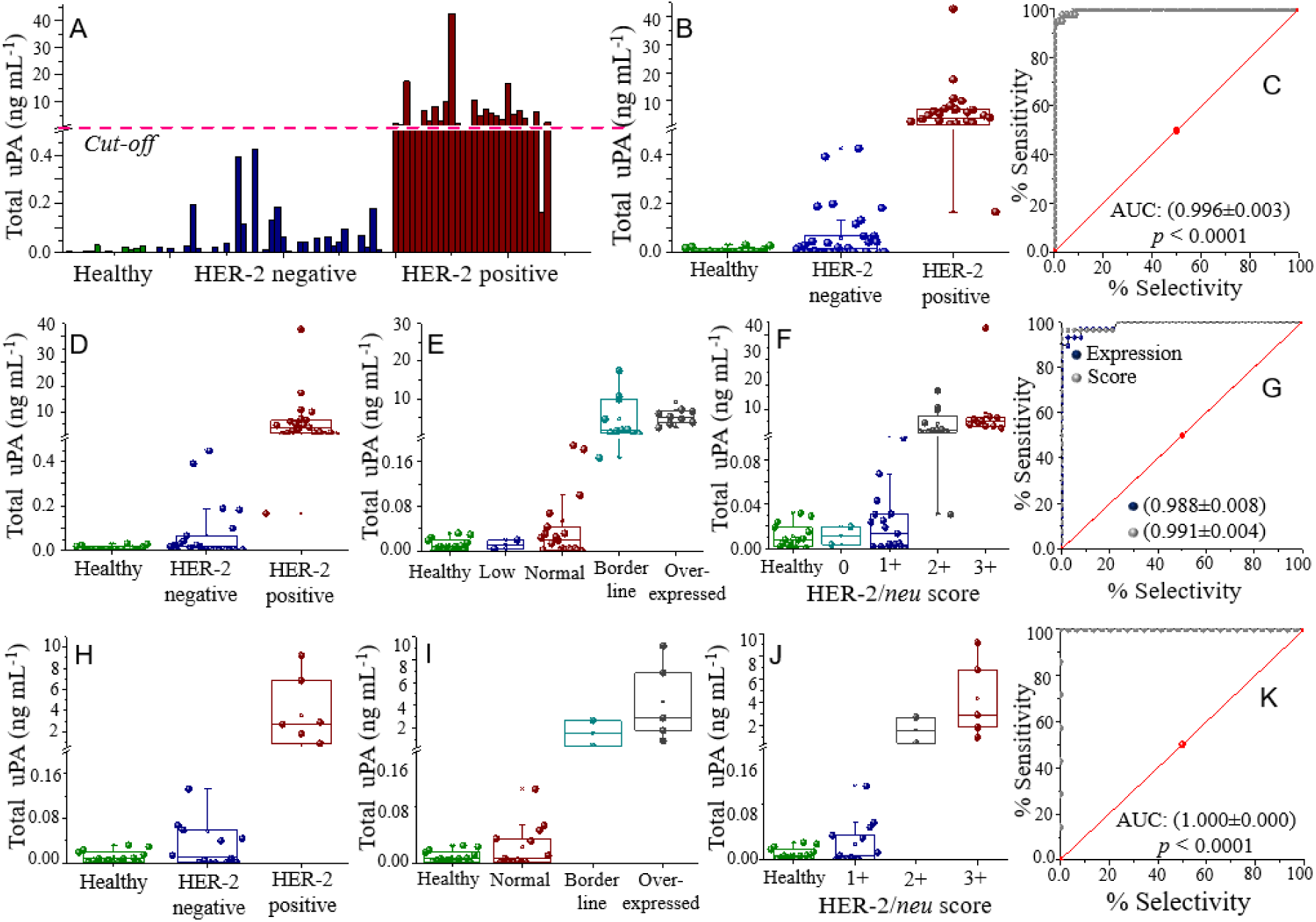
(**A, B**) Correlation between the total serum uPA levels and the patients’ *HER2* status based on analysis of serum samples collected from healthy volunteers (*n* = 15), *HER2*-negative patients (low and normally expressed *HER2* gene) (*n* = 35) and with different *HER2* types (borderline expression and overexpressed *HER2* gene) (*n* = 29). (**C**) ROC curves obtained from the total uPA analysis data. (**D-K**) Segmented analysis of total serum uPA in healthy individuals and *HER2*-negative and allegedly *HER2-*positive cancer patients by cancer type: (**D–G** panel) breast cancer (*n* = 46) and (**H–K** panel) esophagus cancer (*n* = 24). (**D, E, H, I**) Total serum uPA stratified by the *HER2* expression levels, and (**F, J**) by HER-2/*neu* IHC scores. (**G, K**) ROC curves obtained for (**G**) breast and (**K**) esophagus cancer cases based on *HER2* gene expression. All serum samples were diluted 10 times before being examined in triplicates. In all plots, the middle line indicates the median, the error bar means the upper and lower values (according to the 1.5 × IQR criterion, extending to the most extreme non-outlier values), the box refers to the range in which the middle 50% of all data points are, and the upper and lower boxes mean the upper and lower quartiles, which are 25% of the data more or less than those values. Statistical analysis was carried out using Origin 8.5.

ROC analysis of the uPA results (**Figure 1C**) showed a remarkable assay performance, with close to excellent AUC of (0.996±0.003) (*p* < 0.0001). The cut-off value derived was 0.976 ng mL^-1^. The ROC curve’s proximity to the upper-left corner of the ROC space reflected the optimal balance of sensitivity and specificity, confirming accuracy of the *HER2* borderline expression and overexpression discrimination form other cases by serum uPA analysis.

It is important to note that in the 37 patients’ group (borderline expressed and overexpressed *HER2*), 8 patients were treated and supposedly cured to the moment of their serum samples collection synchronized with FDG-PET/CT scanning that showed no tumor present. Their serum uPA results were excluded from the data analysis shown in **Figure 1** and further, being provided in **Figure S3, SI**. Six of these patients had serum uPA levels above the cut-off value.

Total uPA results were stratified by cancer type: breast cancer (*n*=46; **Figures 1D-G**) and esophagus cancer (*n*=24; **Figures 1H-K**). Six cases of gastric and cardia cancer were excluded from comparative analysis of organ specific cancers as they were *HER2*-negative; their serum total uPA was below 0.92 ng mL^-1^ (**Figure S4, SI**). Both allegedly *HER2*-positive specific cancers showed notably high serum uPA levels, on average, (6.843±4.783) ng mL^-1^ (*p* < 0.0001) in breast cancer and (3.546±2.251) ng mL^-1^ (*p* < 0.0001) in esophagus cancer, exceeding 293-fold and 445-fold average uPA concentrations in the serum of *HER2*-negative breast and esophagus cancer patients (**Figures 1D**, **1H**). Thus, elevated serum uPA correlated with a borderline expression and overexpression of *HER2* gene (**Figures 1E**, **1I**). In breast cancer, the borderline expression of *HER2* gene correlated with the average (3.569±2.142) ng mL^-1^ (*p* < 0.0001) serum uPA, increasing to (10.645±8.763) ng mL^-1^ (*p* < 0.0001) in the case of gene overexpression. For comparison, it was just (0.010±0.008) ng mL^-1^ (*p* < 0.001) in serum of patients with normal *HER2* gene expression. In esophagus cancer, the borderline expression correlated with an average (1.583±0.978) ng mL^-1^ serum uPA (*p* < 0.001), and overexpression - with (2.908±1.997) ng mL^-1^ uPA (*p* < 0.0001), with normal *HER2* expression resulting in the meaningly lower serum uPA of (0.008±0.006) ng mL^-1^ (*p* < 0.001). Similarly, the HER-2/*neu* scores 2+ and 3+, reflecting high-density HER-2/*neu* expression on the tumor cell surface, could be linked to the serum uPA concentration higher than found in the serum of cancer patients with tumors characterized by scores 0 and 1+ (considered as *HER2*-negative) (**Figures 1F**, **2J**).

**Figure 2.**
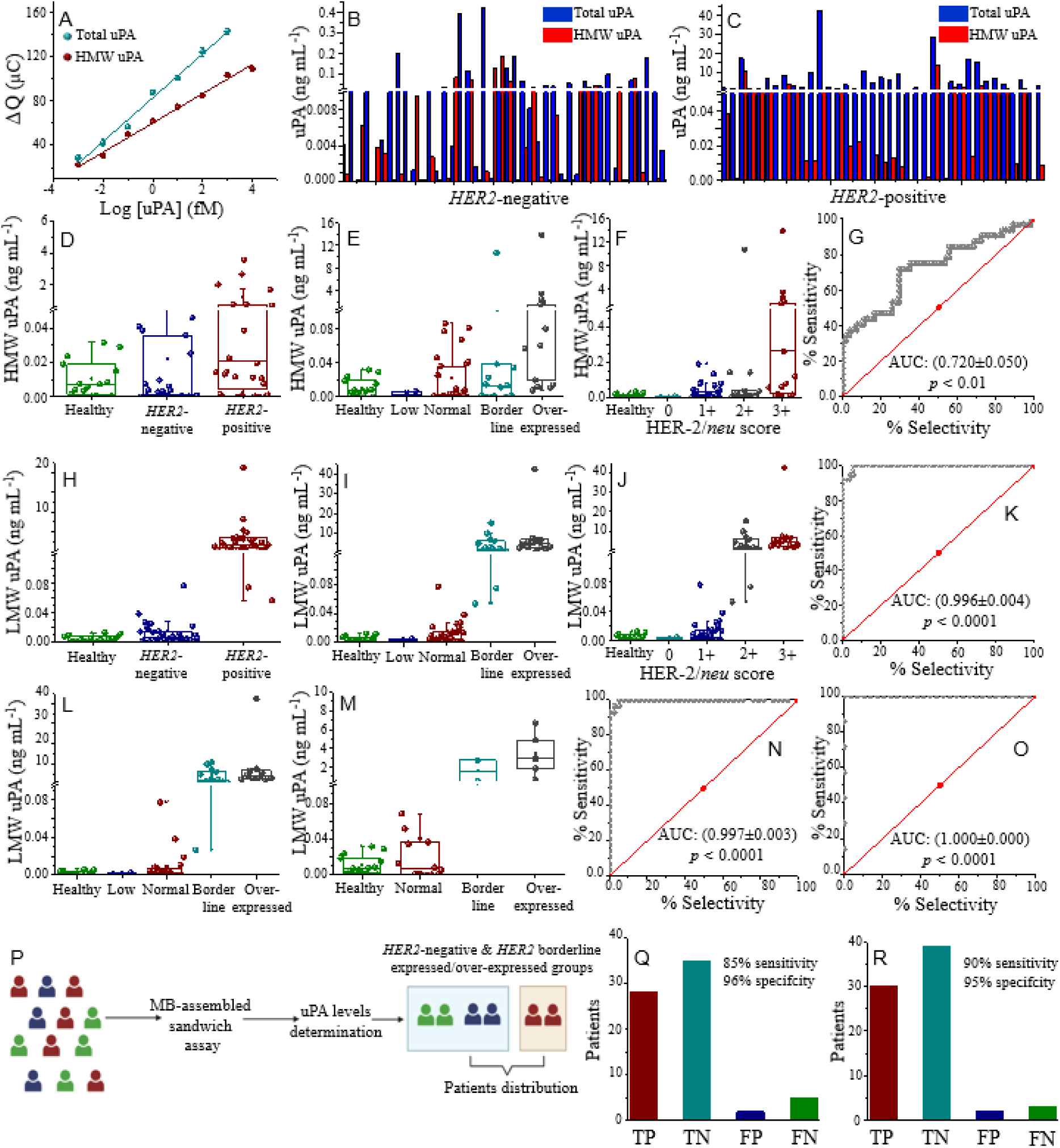
(**A**) The semi-logarithmic dependences of e-ELASA responses on the total uPA (cyan) and HMW uPA (wine) concentrations in uPA-spiked 10% human (error bars are smaller than some concentration dots). (**B, C**) Correlation between the serum total uPA (blue) and HMW uPA (red) levels and the patients’ *HER2* status based on analysis of serum samples from cancer patients with not overexpressed *HER2* gene (*n* = 35) and with allegedly *HER2* positive types (*n* = 29). Healthy volunteers’ data (*n* = 15) are in **Figure S7, SI**. (**D-F**) Serum HMW uPA and (**H-J**) LMW uPA levels stratified by (**D, H**) patients’ cancer status, (**E, I**) *HER2* expression levels, and (**F, J**) HER-2/*neu* ICH scores. (**G, K**) ROC curves obtained for (G) HMW uPA and (K) LMW uPA analysis of tumor’s *HER2* status based on *HER2* gene expression (**L, M**) Serum LMW uPA levels stratified by *HER2* expression levels for (**L**) breast cancer and (**M**) esophageal cancer; and (**N, O**) corresponding ROC curves for (**N**) breast cancer and (**O**) esophageal cancer based on *HER2* gene expression. (**P**) Scheme of patientś liquid biopsy analysis. (**Q, R**) The value of serum uPA analysis for borderline expression and overexpression of *HER2*: true positives (TP), true negatives (TN), false positives (FP) and false negatives (FN) of the allegedly *HER2*-positive group obtained by (**Q**) detecting the total uPA and (**R**) detecting LMW uPA. The sensitivity calculated as TN/(TN+FP) and the selectivity calculated as TP/(TP+FN). The rest notations/conditions are as in Figure 1 (samples preparation and statistics).

ROC curve analysis of serum total uPA data gave quite close cut-off values for *HER2*-positive cancers: 0.892 ng mL⁻¹ uPA for breast cancer (AUC: 0.988±0.008 (*p* < 0.0001)) and 0.867 ng mL⁻¹ uPA for esophageal cancer (AUC: 1.000 ± 0.000) (*p* < 0.0001)) (**Figure 1G, K**). Therewith, slightly better correlations were observed between serum uPA and HER-2/*neu* scores compared to the gene expression data (**Figure 1G**). Considering other parameters, no correlation was found between serum uPA and patients’ sex, weight/height, age, and estrogen receptor status (**Figure S5**, **SI**).

### 3.2. Analysis of specific blood-circulating forms of uPA

In body, uPA exists in several isoforms: its originally expressed pro-uPA zymogen form, high molecular weight uPA (HMW uPA), and low molecular weight uPA (LMW uPA) [24]. In section 3.1, we used aptamers recognizing the sum of all isoforms referred to as total uPA. Yet, in the blood, uPA circulates in two main catalytically active isoforms: HMW uPA and LMW uPA, both may be involved in tumor progression. To determine which form most accurately reflects and/or contributes to *HER2*-associated uPA response, we adapted e-ELASA to selectively detect HMW uPA, by using as a reporter uPA08 aptamer specific solely for HMW uPA [40]. As in total uPA analysis, calibration curves first were constructed for HMW uPA analysis in human serum samples (**Figures 2A** and **S6, SI**). Sensitivity of HMW uPA detection in serum, 12.912 µC fM^-1^, was close to that in PBS, and the LOD was 1 aM (by IUPAC definition) and 0.430 aM (as previously specified). The calibration curves linear equations and sensitivities of analysis were:

Total uPA analysis in PBS: *y* = 20.28*x* + 83.87, *R*² = 0.99 (the sensitivity: 20.28 ± 5.21 μC fM^-1^), and in 10% serum: *y* = 19.14*x* + 82.93, *R*² = 0.99 (the sensitivity: 19.14 ± 4.50 μC fM^-1^). HMW uPA analysis in PBS: *y* = 13.40*x* + 63.96, *R*² = 0.99 (the sensitivity: 13.40 ± 3.23 μC fM^-1^), and 10% serum: *y* = 12.91*x* + 60.93, *R*² = 0.99 (the sensitivity: 12.91 ± 3.03 μC fM^-1^).

Both serum HMW uPA and LMW uPA levels were analyzed in serum samples of patients with tumors showing borderline expression of *HER2* and in *HER2*-positive tumors, by using the total uPA and HMW uPA specific e-ELASAs. LMW uPA was calculated by subtracting the detected HMW uPA concentration from the total uPA concentration. This dual assessment provided deeper insights into serum uPA profiles in patients with different HER-2/*neu* tumor statuses. We found that serum HMW uPA did not show clear variation patterns as total uPA did, indicating that serum HMW uPA contributed weakly to the accuracy of *HER2*-associated cancer status determination (**Figures 2B-G)**. Although median HMW uPA levels were higher in the groups with overexpressed (0.801 ng mL^-1^; IQR: 0.001 ng mL^-1^ – 1.617 ng mL^-1^; *p* < 0.09) and borderline expressed *HER2* (0.013 ng mL^-1^; IQR: 0.002 ng mL^-1^ – 0.038 ng mL^-1^; *p* < 0.48) than in *HER2*-negative (0.023 ng mL^-1^; IQR: 0.0002 ng mL^-1^ – 0.0350 ng mL^-1^; *p* < 0.31) and healthy individuals (0.011 ng mL^-1^; IQR: 0.001 ng mL^-1^ – 0.018 ng mL^-1^; *p* < 0.5) groups, the differences were not statistically significant. The standard deviations were too large, and HMW uPA values overlapped largely across all groups (**Figure 2D-F**). A very weak (if any) correlation was found between serum HMW uPA and tumor status, *HER2* expression, and HER-2/*neu* scores. The ROC analysis yielded an AUC of 0.720 ± 0.050 (*p* < 0.05), indicating poor ability of HMW uPA to stratify *HER2* status of tumors.

In contrast, serum LMW uPA correlated remarkably well both with *HER2* expression levels and HER-2/*neu* scores (**Figure 2H-J**). Notably, LMW uPA constituted the major fraction of total uPA, and concentrations of both forms showed strong correlations across all studied groups, in contrast to HMW uPA (**Figure S8, SI**). Serum LMW uPA levels’ average was statistically significantly higher in patients with *HER2* overexpression (6.447±4.722) ng mL^-1^ (*p* < 0.0001) and with borderline expression (3.105±2.044) ng mL^-1^ (*p* < 0.0001) than in *HER2*-negative (0.042±0.030) ng mL^-1^ (*p* < 0.001) and healthy individuals (0.011±0.009) ng mL^-1^ (*p* < 0.001) groups. ROC curve analysis gave an AUC of 0.996 ± 0.004 (*p* < 0.0001) supporting the diagnostic utility of LMW uPA as a biomarker for *HER*2-borderline expressed and *HER*2-overexpressed cancers, with a cut-off value of 0.917 ng mL^-1^.

The accuracy of *HER2*-borderline expressed and *HER2*-overexpressed cancer detection by serum LMW uPA analysis remained similarly high across different cancer types (**Figure 2L, M**). In both breast and esophageal cancer cohorts, the average LMW uPA concentrations were markedly elevated in *HER2*-overexpressed ((6.267±4.023) ng mL^-1^; *p* < 0.0001 for breast cancer, and (3.367±2.415) ng mL^-1^; *p* < 0.0001 for esophageal cancer) and borderline expressed (3.426±1.963) ng mL^-1^; *p* < 0.0001 for breast cancer, and (1.583±1.122) ng mL^-1^; *p* < 0.0001 for esophageal cancer) cases compared to *HER2*-negative cases ((0.044±0.010) ng mL^-1^; *p* < 0.0001, and (0.041±0.006) ng mL^-1^; *p* < 0.0001, for breast and esophageal cancer, respectively). These differences were statistically significant and enabled clear distinctions between *HER2*-negative and allegedly *HER2*-positive cancers, with no overlap between groups. ROC curve analysis yielded exceptional AUCs of 0.997±0.003 (*p* < 0.0001), for breast cancer (**Figure 2N**), and 1.000±0.000 (*p* < 0.0001), for esophageal cancer (**Figure 2O**), and cut-offs for allegedly *HER2*-positive cancers of 0.917 ng mL^-1^ LMW uPA for the overall cancer dataset, 0.857 ng mL^-1^ for breast cancer, and 0.825 ng mL^-1^ for esophageal cancer.

Thus, serum total uPA correctly identified 28 out of 29 patients with *HER2*-positive tumors and tumors with borderline expressed *HER2* as true positives, with 1 case classified as false positive, resulting in the cancer stratification accuracy of 96.6% (**Figure 2P, Q**). Serum LMW uPA allowed 93.1% accurate discrimination of *HER2*-positive tumors and tumors with borderline expressed *HER2* (**Figure 2P, R**).

## 4. Discussion

Our data show that serum total uPA informs 96.6% accurately about the *HER2* tumor status (*HER2* borderline expression and overexpression) and thus enables these cancer subtypes stratification. Yet, serum uPA levels in patients with cancers characterized by *HER2* borderline expression (including both *HER2*-negative and positive cancers) and overexpression (*HER2*-positive cancers) showed similar patterns and cannot be distinguished. The total uPA concentration above 0.974 ng mL^-1^ (the cut-off value) informed correctly about the *HER2* expression status of tumors (borderline expressed or overexpressed) in 29 patients, with one patient giving a false negative result. Serum uPA informed about *HER2* expression levels independently of such parameters as patients’ sex, weight/height, age, and estrogen receptor status. An important observation is that six of the eight diagnosed *HER*2–positive cancer patients with no PET-detectable tumors at the time of serum collection and analysis still had high serum uPA levels (**Figure S3, SI**). As we lack sufficient data to determine whether these cases represent true remission or relapse, we simply acknowledge this observation. We can speculate, however, that uPA overexpression may occur independently of anti-*HER2* treatment, and that the established prognostic role of serum uPA might be reflected in its elevated levels in patients considered to be in remission. Both hypotheses require verification in a larger cohort of treated patients with longitudinal serum analyses.

Therewith, analysis of different forms of uPA, HMW uPA and LMW uPA, shows that LMW uPA and not HMW uPA correctly informs about the *HER*2 status of tumors. When contribution of serum HMW uPA to total uPA is disregarded, serum LMW uPA informs 95% specifically about *HER2* expression status in all types of cancers studied (breast, esophagus, gastric, and cardia cancers, **Figure 2H-K**), with a *HER2* borderline expression and *HER2*-positive cancer cut-off of 0.917 ng mL^-1^. Both breast cancer and esophagus cancer cut-offs showed close values.

Another important observation is that serum LMW uPA and total uPA levels in healthy individuals and in *HER2*-negative breast, esophagus, gastric and cardia carcinomas and adenocarcinomas are not so different, though the median value was slightly higher in cancer patients with normally expressed *HER2* and with HER-2/*neu* score 1+ (**Figure 2J**). Thus, serum LMW uPA did not inform generally about cancer. Our results underscore the strong discriminatory capacity of serum uPA to differentiate specifically between *HER2*-negative (score 0 and 1+) and alleged and true *HER2*-positive conditions (scores 2+ and 3+). However, they also raise questions about the underlying pathophysiological processes.

Normally, uPA converts plasminogen to plasmin, a broad-spectrum serine protease that promotes ECM degradation both directly and indirectly, via activation of matrix metalloproteinases, contributing to fibrinolysis by degrading fibrin, the main protein component of blood clots, and some other ECM components [24,44]. In a healthy organism, plasmin is active in a variety of tissue-remodeling processes helping preventing blood clots, thrombosis, cardiovascular diseases, combatting inflammation and ensure proper wound healing [45]. Yet, in cancer, activation of plasminogen by uPA not only assists proteolytic degradation of ECM, but also causes several pathological processes contributing to tumorigenesis, tumor invasion and metastasis [44]. The reasons for emerging pathophysiological routes are not completely clear, as well as the role played by different catalytically active forms of uPA.

uPA is secreted as a single-chain catalytically-inactive zymogen form, referred to as pro-uPA, comprising three distinct domains: the growth factor domain (GFD), the kringle domain (KD), and the catalytic serine protease domain (**Figure 3A**) [24,44]. Pro-uPA is then proteolytic cleaved and converted, reportedly by other proteases such as plasmin, trypsin, kallikrein, cathepsin B, human T cell associated serine proteinase-1, and thermolysin [24], into catalytically active double-chain HMW uPA, in which the GFD-KD region (an A chain, involved in binding to receptors and substrates) remains connected to the catalytic domain (a B chain responsible for its enzymatic activity) via a disulfide bond (**Figure 3A**).

**Figure 3.**
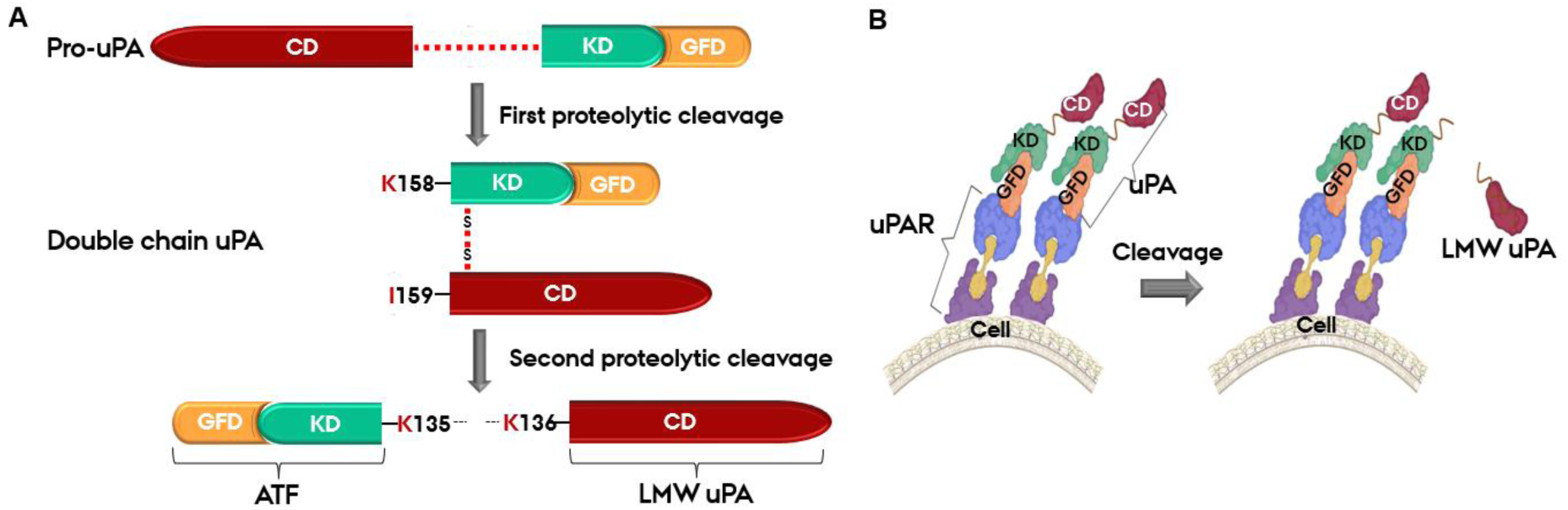
(**A**) Schematic representation of the uPA activation and proteolytic cleavage. The single-chain precursor form, pro-uPA, contains 3 structural domains: the growth factor domain (GFD), kringle domain (KD), and catalytic domain (CD). Activation of pro-uPA occurs via cleavage at the Lys158–Ile159 peptide bond, producing the two-chain uPA held together by a disulfide bridge (HMW uPA, ca. 54 kDa). 2^nd^ proteolytic cleavage between Lys135 and Lys136 yields two fragments: the amino-terminal fragment (ATF), including GFD-KD, and the catalytic LMW form (LMW uPA, ca. 33 kDa), comprising the serine protease domain. Adapted from [24]. (**B**) uPA binding to uPAR dimers on the cell surface facilitates 2^nd^ proteolytic cleavage leading to LMW uPA release into the bloodstream.

Further proteolytic cleavage of HMW uPA produces the amino terminal fragment, ATF, comprising the GFD-KD region, capable of binding to uPAR, and a soluble LMW form, which is the catalytic domain and the most part of the structurally disordered 27-residue linker, via which it was tethered to ATF (**Figure 3**). This cleavage happens extracellularly, supposedly after uPA binds to its receptor uPAR on the cell surface, although the exact mechanism remains unclear [46]. Binding of both pro-uPA and HMW uPA to the cell-surface anchored uPAR activates surface conversion of plasminogen to plasmin and may indeed trigger the pathophysiological pathways of tumor development [24].

There are two questions concerning the relation between serum uPA and *HER2*-positive cancers:

1). Why such proteolytic enzyme as uPA is co-overexpressed with *HER2* [26,47]?

2). Why does LMW uPA and not HMW uPA consistently elevate in the blood of patients with *HER2*-positive tumors and tumors with borderline-expressed *HER2*?

Regarding question (1), it was suggested that in *HER2-*positive cancers activation of *HER2* induced strong signaling via the protein kinase Cα and steroid receptor co-activator, both being critical components of the uPA/uPAR-mediated cancer cell invasion, or nuclear factor-kappaB, capable of mediating HER-2/*neu* and uPAR expression in cancer stem cells [25]. This could lead to activation of ETS and Kruppel-like family of transcription factors, whose binding to the promoter regions of *HER2* or *PLAUR* (gene symbol for uPAR) gene then leads to their co-amplification and consequent overexpression of HER-2/*neu* and both uPAR and uPA, since the latter two strongly correlate [25].

Regarding question (2), LMW uPA cannot bind to uPAR and remains soluble, while still retaining full catalytic activity. When the uPA/uPAR system is overexpressed, a proportionally greater fraction of LMW uPA in total uPA is then expected to freely circulate in the blood. Pro-uPA and HMW uPA are predominantly cell-bound forms and should therefore constitute a proportionally smaller fraction of serum total uPA. Yet, why is LMW uPA (and not HMW uPA) elevated? We suggest it is closely connected with the way plasmin-uPA system is activated on the cell surface. By cleaving and activating each other, uPA and plasmin establish a positive feedback loop. It is debated how the whole catalytic plasmin-uPA activation circle starts. Recent *in vitro* studies show that HMW uPA can both activate pro-uPA and undergo autocatalytic self-cleavage releasing LMW uPA, an activity that plasmin cannot perform [46]. Then, the autocatalytic cleavage of HMW uPA may be the most plausible mechanism for the LMW uPA shedding into the bloodstream, since:

(i). Several studies report on uPAR dimerization on the cell surface and uPA binding to uPAR dimers [46,48,49]. Overexpression of uPA/uPAR in *HER2*-positive tumors then results in high-density uPA/uPAR dimer complexes on the cell surface.

(ii). Binding of uPA to the dimers brings two HMW uPA (or pro-uPA) molecules capable of autoactivation in the absence of plasmin [46] in close proximity, which promotes autocatalytic proteolytic cleavage of uPA releasing LMW uPA (**Figure 3B**). Thus, uPA/uPAR overexpression in *HER2*-positive tumors can lead indeed to higher levels of LMW uPA in the blood of *HER2*-positive cancer patients. As follows from our data, borderline overexpression of *HER2* can also trigger overexpression of uPA and its release in the bloodstream.

We compared our findings with previously published data. Most studies focus on uPA analysis in tumors and link elevated tumor uPA with higher disease risk and poorer prognosis [23,24,30,33,50–56]. Serum uPA state/status may inherently differ from those in tissues yet provide an alternative modality for tumor assessment, via liquid biopsy. However, liquid biopsy studies are limited and primarily focused on prognostic significance of serum uPA, examining connections between serum total uPA and cancer survival outcomes [36,57–60].

In head and neck squamous cell carcinomas, the median concentration of serum pro-uPA and HMW uPA was 0.48 ng mL^-1^ (range: 0.24–1.92 ng mL^-1^, *p*-value “*not specified*”), with no significant difference between healthy volunteers (*n*=28) and cancer patients (*n*=40) [57]. The data exhibited high variability, interfering with meaningful statistical analysis. In anoter study, elevated serum uPAR, rather than total uPA (median: 0.69 ng mL^-1^; range: 0-4.76 ng mL^-1^), was linked to an increased risk of soft-tissue sarcoma-related mortality, with no correlation between serum uPA and clinicopathological parameters (*n*=79) [58]. Patients with pancreatic ductal adenocarcinoma showed a mean serum uPA level of 3.23±1.84 ng mL^-1^ (range: 1.24–7.6 ng mL^-1^; median: 2.75 ng mL^-1^), compared to 2.18±1.45 ng mL^-1^ in chronic pancreatitis patients (range: 0.88–5.4 ng mL^-1^; median: 1.51 ng mL^-1^) and 1.01±0.32 ng mL^-1^ in the control group (range: 0.2-1.66 ng mL^-1^; median: 1.05 ng mL^-1^). Although the differences among the three groups were statistically significant (*p* < 0.01), the diagnostic accuracy was limited due to the substantial overlap of the results and high variability in serum uPA analysis [60]. A significant correlation was reported between elevated serum uPA (>2 ng mL^-1^) and reduced patients’ survival (181±155 days vs. 335±314 days for < 2 ng mL^-1^ uPA; *r* = −0.391; *p* < 0.05) [60]. These results also exhibited high variability. A serum uPA cut-off of 1 ng mL^-1^ predicted hepatocellular carcinoma-related death with a 41% sensitivity and 77.5% specificity (*n*=287, *p* < 0.002) [59]. The median serum uPA concentration was reported as 0.7 ng mL^-1^ (range 0.2–14.7 ng mL^-1^), or as a mean: 1.0±1.36 ng mL^-1^. In a study of metastatic breast cancer patients (*n*=252), a serum uPA cut-off value of 2.52 ng mL^-1^ was associated with shorter overall and progression-free survival (*p* < 0.001) and the presence of visceral metastases and multiple metastatic sites [36]. High variability in reported results may stem from uncertainties in the quality of ELISA kits employed for analysis [36,58–60], currently unavailable on the market, results being often compromised by inconsistent methodological details provided. More accurate analytical tools might yield more accurate data linking serum uPA and cancer statuses.

Our and other findings point at the existence of two serum uPA patterns in cancer: elevated uPA levels linked to either (1) poor prognosis and metastatic disease or (2) *HER2*-associated cancers that may be also extended to other cancer subtypes driven by similar pathophysiological mechanisms. Therewith, LMW uPA, dominating in the blood of cancer patients, may play a significant role in cancer progression due to the loss of regulatory domains and persistence of unregulated protease activity. At its elevated levels, LMW uPA may lead to diffuse-enhanced ECM degradation, promoting invasion and metastasis, and contribute to more aggressive tumor behavior through uncontrolled ECM remodeling and activation of other proteases, such as matrix metalloproteinases via plasmin. Lastly, uPAR not only anchors HMW uPA and pro-uPA but also mediates signaling via integrins, EGFR, and other pathways. LMW uPA bypasses this axis, as it does not engage uPAR-mediated signaling involved in processes such as cell survival and migration. However, its proteolytic role continues, potentially contributing to angiogenesis, immune evasion, and chemoresistance through ECM remodeling.

To conclude, we have shown that liquid biopsy testing of serum uPA enables stratification of *HER2*-positive cancers and *HER2*-borderline expressed cancers. In 85 patients’ trial, with 15 healthy individuals as a control group, serum total uPA exceeding 0.976 ng mL^-1^ 96.6% accurately informed about *HER2* expression levels in the HER-2/*neu* score 2+/3+ patients’ group. This new classification of allegedly *HER2*-positive cancers, based on serum uPA, could help identify the right patients for treatment yet needs further evaluation in the future.

## Additional Information

## Acknowledgements

We acknowledge Novo Nordisk Foundation for funding of the project “Validating Serum Tests for Human Epidermal growth factor Receptor-2 for Precise Diagnosis and Stratification of Breast and Gastro-Oesophageal Cancers”, grant reference number NNF20OC0065428, and the European Union’s Horizon research and innovation program under the Marie Skłodowska-Curie HORIZON – MSCA – 2023 – DN Grant Agreement No. 101169504. We thank Assoc. Prof. Trine Tramm, Institute of Clinical Medicine and Pathology, Aarhus University, for critically reading and commenting on our work.

## Author contributions

M.E.J.L.M. and S.B.: Investigation; Formal analysis; Data curation, Visualization, Validation; Writing-Original draft preparation; N.L.C.: Patients’ samples collection; Summary of pathology reports; M.A.P.: Samples’ collection and processing protocol and clinical design; K.R.J.: Image analysis; Patients stratification; M.H.V.: Image collection and analysis; Formal analysis; Data curation; Reviewing and Editing; Funding acquisition; E.E.F: Conceptualization, Supervision, Formal analysis; Hypothesis validation; Results elaboration; Writing-Original draft preparation, Reviewing and Editing; Funding acquisition.

## Ethics approval and consent to participate

Blood samples from healthy volunteers and patients FDG-PET/CT scanned on suspicion of breast, esophagus and gastric cancers were collected at Aarhus University Hospital, Aarhus, Denmark. All subjects signed an informed consent form. The study was approved by the Regional Ethics Committee for Science in the Central Region of Jutland, Denmark, approval number 1-10-72-168-22. The performed research fully complies with the Helsinki Declaration (in its latest versions).

## Supporting Information

List of abbreviations used; Raw analytical data and calibration curves; Additional information on statistical analysis and data processing.

## Data availability

Data supporting the results reported in the article can be found in the SI files.

